# Association between psychological distress trajectories from adolescence to midlife and mental health during the pandemic: Evidence from two British birth cohorts

**DOI:** 10.1101/2021.09.30.21264342

**Authors:** Vanessa Moulton, Alice Sullivan, Praveetha Patalay, Emla Fitzsimons, Morag Henderson, David Bann, George B. Ploubidis

## Abstract

**Background:** Some studies suggest worsening mental health in the early stages of the pandemic, for individuals with pre-existing mental health conditions the evidence is mixed. We examined whether different life-course trajectories of psychological distress from adolescence to midlife were associated with psychological distress, lower life satisfaction and feelings of loneliness at different stages during the pandemic.

**Methods:** This study is a secondary analysis of two nationally representative British Birth cohorts, the National Child Development Study (1958) and 1970 British Cohort Study, from birth to later mid-life. We used latent variable mixture models to identify pre-pandemic longitudinal trajectories of psychological distress and a modified poisson model with robust standard errors to estimate associations with mental health outcomes during the pandemic from May 2020 to March 2021.

**Findings:** Our analysis identified five distinct pre-pandemic trajectories of psychological distress in both cohorts. All trajectories with prior symptoms of psychological distress were associated with a greater relative risk of mental health outcomes during the pandemic. This was the case irrespective of age of onset, severity, longevity and proximal occurrence. Those who had experienced more than one prior episode of high psychological distress, and more recent occurrences, faced the greatest risk of poor mental health during the pandemic.

**Interpretation:** Whilst any prior episode of poor mental health put individuals at greater risk of severe mental health symptoms, those with chronic and more recent occurrence are likely to require greater mental health support.

## Introduction

The COVID-19 pandemic and accompanying policy measures may have affected population mental health. In the early stages of the pandemic, some studies showed significantly increased levels of poor mental health.^1,2,3,5^. For individuals with pre-existing mental health conditions the evidence is mixed, some studies suggest worsening mental health in the early stages of the pandemic^3,5^, while other studies show no increase in symptoms^2,4^ and a graded dose-response relation based on prior number of symptoms and chronicity.^4^ Many studies have employed retrospective measures of pre-existing mental health.^5^ Even in prospective studies, pre-COVID-19 mental health data has typically been measured at one time point only or over the short-term.^3^ Few studies have examined the relation of pre-existing mental health over the longer term^2,4^.

No extant studies have investigated life-course trajectories of mental health in the general population prior to the COVID-19 outbreak.^6,7^ The risk of poor mental health in the general population has been shown to be heterogenous, following different longitudinal trajectories that vary in terms of age of onset, symptom severity and risks of recurrence.^8,9^ Recent studies of mental health during COVID-19 have identified heterogenous trajectories of mental health during the first six months of the pandemic.^10^ Distinct pre-pandemic life-course mental health trajectories based on onset, severity and stability across the adult life-span may also be related to poor mental health during the pandemic.

Mental well-being is distinct from mental illness, and predicts important outcomes including physical health. Subjective wellbeing is sometimes favored by governments as positive measures are deemed better for evaluating public health approaches.^11,12^ The pandemic and measures used to mitigate the crisis may have a disproportionate influence on those with mental health difficulties because of increased isolation and loss of social connectedness. ^13^ Loneliness is a key predictor of mental health challenges particularly in older adults. ^14^ We examine trajectories of mental health across the life-course from adolescence to midlife, thus giving an overview of the timing of onset (adolescence, early or mid-adulthood), along with the severity and stability at different life-stages.

We do so by exploiting the rich longitudinal data available in two large nationally representative British birth cohorts from birth and across the life-course to investigate whether, i) differing psychological distress trajectories in these populations were more likely to result in symptoms of depression and anxiety, lower life satisfaction and feelings of loneliness during the COVID-19 pandemic, and ii) whether there were differences in these mental health outcomes in later mid-life, for life-course psychological distress groupings at three-distinct timepoints, from May 2020 through to March 2021, as the pandemic developed.

## Methods

### Participants

Our data are from two ongoing population-based birth cohorts:

#### 1958 National Child Development Study (NCDS)

The NCDS follows the lives of 17,415 people that were born in England, Scotland or Wales in a single week in March 1958. The NCDS started as the Perinatal Mortality Survey and captured 98% of the total births in Great Britain in the target week. The cohort has been followed up eleven times between ages 7 and 55.^15^

#### 1970 British Cohort Study (BCS70)

The BCS70 follows the lives of 17,198 people (representing 95% to 98% of the target population) born in England, Scotland and Wales in a single week in April 1970. Participants have since been followed up nine times between ages 5 and 46.^16^

In addition, during the COVID-19 pandemic participants of the NCDS and BCS70 completed an online survey at three different time-points when they were aged 62 and 50, respectively. The first survey was conducted during the first national lockdown, between 4 and 26 May 2020 (Wave 1: NCDS N: 5,178; BCS70 N: 4,223). The second survey was completed between 10 September and 16 October 2020 (Wave 2: NCDS N: 6,282; BCS70 N: 5,320) when the first national lockdown had been lifted, but restrictions on social contact still remained, and the third survey was conducted during the third national lockdown, between 1 February and 21 March 2021 (Wave 3: NCDS N: 6,757; BCS70 N:5,684).^17^

Our analytic sample includes all participants in the NCDS and BCS70 surveys, excluding those who had died or emigrated by age 50 in the NCDS and age 46 in the BCS70. The sample size for NCDS is n=15,291 and for BCS70 n=17,486 (sample descriptive statistics are presented in supplementary Table S1). To deal with attrition and item non-response and to restore sample representativeness we used Multiple Imputation (MI) with chained equations, imputing 25 data sets at each wave.^18^ All variables used in our main analysis, as well as a set of auxiliary variables were included in the imputation models to maximise the plausibility of the ‘missing at random’ (MAR) assumption in order to reduce bias due to missing data.^19^ As an additional sensitivity analysis all models were rerun in line with the ‘impute and delete’ method and resulted in similar findings (available in supplementary Table S2).

### Measures

#### Pre-pandemic psychological distress

Psychological distress was measured in both cohorts with the nine-item version of the Malaise Inventory ^20^ from ages 23, 33, 42 and 50 in the NCDS and ages 26, 34, 42 and 46 in the BCS70. Psychological distress captures depression and anxiety symptoms. In both surveys the Malaise items were assessed via written self-completion, either on paper or via computer. The Malaise inventory has been shown to have good psychometric properties,^21^ measurement invariance, and has been used in general population studies as well as investigations of high risk groups.^22^ In both cohorts, at age 16 four items from the Children’s Behavior Questionnaire (CBQ) reflective of affective disorders (Low mood, irritability, worry, and fearfulness), as reported by the child’s mother were employed.

#### Outcomes

There were three outcome measures, psychological distress, life satisfaction and loneliness, completed at three-time points during the pandemic. *Psychological distress* was measured in both cohorts with the nine-item version of the Malaise Inventory,^20^ the same measure assessed across the life-course pre-pandemic. The measure was transformed into a dichotomous variable where 1 represents high psychological distress (rating ≥4). Subjective life satisfaction was measured by asking ‘Overall, how satisfied are you with your life nowadays, where 0 means ‘not at all’ and 10 means ‘completely’?’. Research has shown that single item measures of life satisfaction are highly correlated with longer life satisfaction scales.^23^ The measure was transformed into a dichotomous variable where 1 is high or very high level of satisfaction (rating ≥ 7). Loneliness was measured using the three-item short form of the Revised UCLA loneliness scale. Items were assessed on a three-point scale to produce a loneliness score from 3 to 9, a dichotomous variable was created using a cut-off score of 6 to represent presence of loneliness.^24^ As sensitivity analysis we estimated models with continuous versions of all outcomes with appropriate link functions and transformations where possible (available in supplementary Table S6).

#### Potential confounders

We include in our analysis a rich set of variables comprising early life factors (sex, ever breastfed, mother smoked daily during pregnancy, gestation period, and birthweight), socio-economic factors (parental social class, education, housing tenure, access to amenities, crowding and marital status), parental factors (maternal age at birth, mother worked at all in first five years, separated from child and read to), child behavior and health (cohort member bedwetting since age 5, had any medical conditions, and Body Mass Index (BMI)), and cognitive ability (details are available in supplementary Table S3).

### Analytic approach

As a first step, we used latent variable mixture models to identify longitudinal typologies of mental health^8^ employing psychological distress at five time points from adolescence to mid-life.^20^ We applied a four-category ordinal variable on the basis of grouping factor scores distributions of the latent measures (from the 1^st^ to 50^th,^ 51^st^ to the 75^th^, 76^th^ to 90^th^ and the 91^st^ to 100^th^ percentile) to each of the five time points in each cohort. (Further details are available in supplementary Tables S4a and S4b).

We first examined the proportion of individuals in each of the mental health classes prior to the pandemic with psychological distress, life-satisfaction, and loneliness during the pandemic. We used a modified Poisson model with robust standard errors that returns risk ratios for ease of interpretation and to avoid bias due to non-collapsibility of the odds ratio^25^ to estimate associations between the psychological distress trajectories and mental health outcomes during the pandemic.

To aid interpretation we present between trajectories comparisons of marginal effects on psychological distress, life satisfaction and loneliness at three-time points during the pandemic, thereby answering question 2. All Latent mixture modelling was conducted using MPLUS v8.2, and descriptive analyses, GLM Poisson regression and multiple imputation were done using Stata version 15 (StataCorp).

## Results

### Trajectories of psychological distress from adolescence to midlife

In both cohorts we identified five longitudinal groups (latent classes) of psychological distress as the most parsimonious models (Table S4a and S4b in the supplement). Figure 1a and 1b shows the means for each longitudinal latent class on each of the five measures in the NCDS (age 16 to 50) and BCS70 (age 16 to 46).

**Figure 1a:**
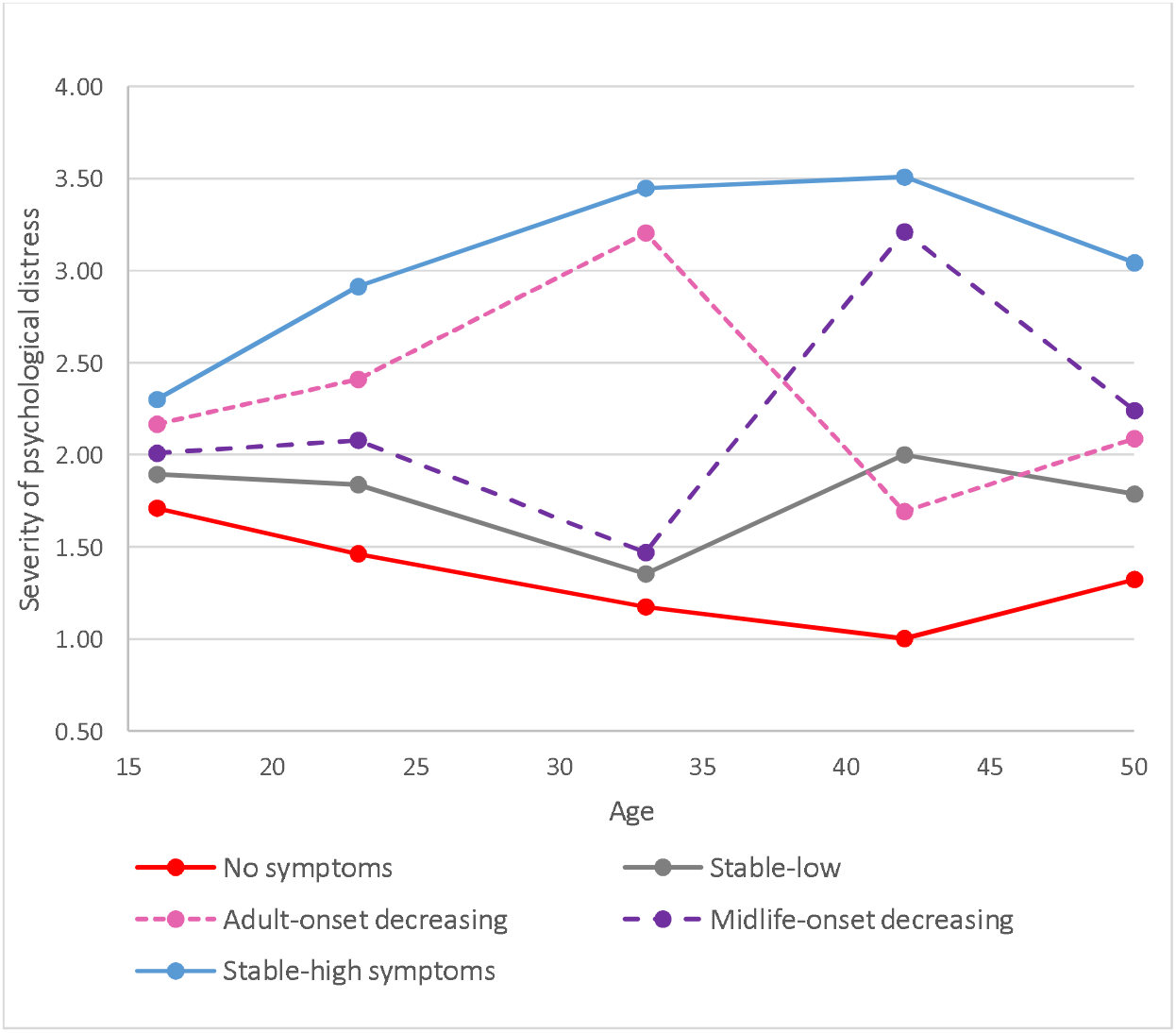
Five longitudinal classes of psychological distress from age 16 to 50 in the NCDS.

**Figure 1b:**
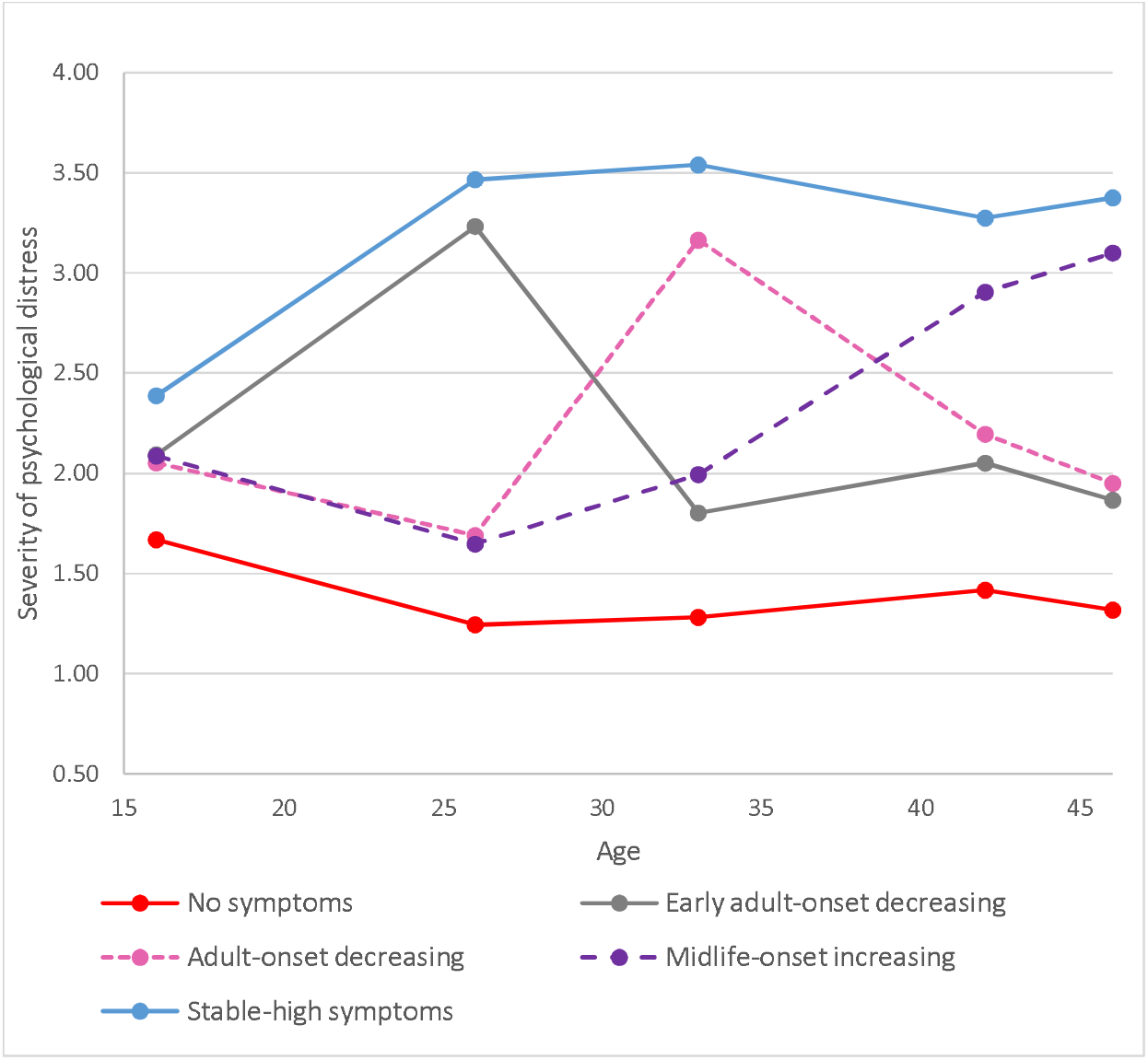
Five longitudinal classes of psychological distress from age 16 to 46 in the BCS70.

Descriptive statistics of the trajectories of psychological distress and mental health outcomes during the pandemic are presented in Table 1. The five longitudinal groups were similar, but not identical in the two cohorts.

**Table 1:**
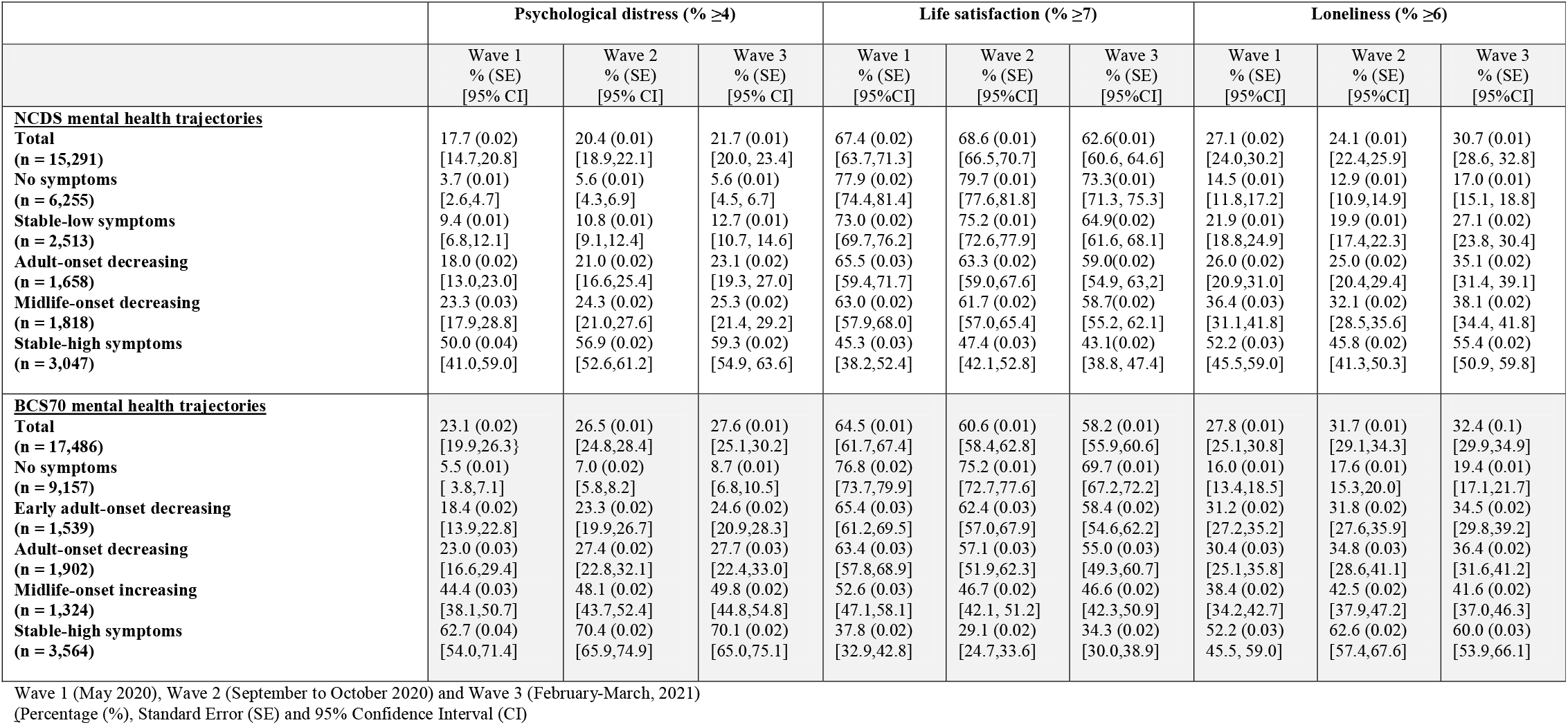
**Percentage of cohort members with high psychological distress, high life satisfaction, and feelings of loneliness by pre-pandemic psychological distress trajectories in the NCDS and BCS70 during the COVID-19 pandemic**

|  | Psychological distress (% ≥4) |  |  | Life satisfaction (% ≥7) |  |  | Loneliness (% ≥6) |  |  |
| --- | --- | --- | --- | --- | --- | --- | --- | --- | --- |
|  | Wave 1<br>% (SE)<br>[95% CI] | Wave 2<br>% (SE)<br>[95% CI] | Wave 3<br>% (SE)<br>[95% CI] | Wave 1<br>% (SE)<br>[95% CI] | Wave 2<br>% (SE)<br>[95% CI] | Wave 3<br>% (SE)<br>[95% CI] | Wave 1<br>% (SE)<br>[95% CI] | Wave 2<br>% (SE)<br>[95% CI] | Wave 3<br>% (SE)<br>[95% CI] |
| **<u>NCDS mental health trajectories</u>** |  |  |  |  |  |  |  |  |  |
| **Total**<br>**(n = 15,291)** | 17.7 (0.02)<br>[14.7,20.8] | 20.4 (0.01)<br>[18.9,22.1] | 21.7 (0.01)<br>[20.0, 23.4] | 67.4 (0.02)<br>[63.7,71.3] | 68.6 (0.01)<br>[66.5,70.7] | 62.6(0.01)<br>[60.6, 64.6] | 27.1 (0.02)<br>[24.0,30.2] | 24.1 (0.01)<br>[22.4,25.9] | 30.7 (0.01)<br>[28.6, 32.8] |
| **No symptoms**<br>**(n = 6,255)** | 3.7 (0.01)<br>[2.6,4.7] | 5.6 (0.01)<br>[4.3,6.9] | 5.6 (0.01)<br>[4.5, 6.7] | 77.9 (0.02)<br>[74.4,81.4] | 79.7 (0.01)<br>[77.6,81.8] | 73.3(0.01)<br>[71.3, 75.3] | 14.5 (0.01)<br>[11.8,17.2] | 12.9 (0.01)<br>[10.9,14.9] | 17.0 (0.01)<br>[15.1, 18.8] |
| **Stable-low symptoms**<br>**(n = 2,513)** | 9.4 (0.01)<br>[6.8,12.1] | 10.8 (0.01)<br>[9.1,12.4] | 12.7 (0.01)<br>[10.7, 14.6] | 73.0 (0.02)<br>[69.7,76.2] | 75.2 (0.01)<br>[72.6,77.9] | 64.9(0.02)<br>[61.6, 68.1] | 21.9 (0.01)<br>[18.8,24.9] | 19.9 (0.01)<br>[17.4,22.3] | 27.1 (0.02)<br>[23.8, 30.4] |
| **Adult-onset decreasing**<br>**(n = 1,658)** | 18.0 (0.02)<br>[13.0,23.0] | 21.0 (0.02)<br>[16.6,25.4] | 23.1 (0.02)<br>[19.3, 27.0] | 65.5 (0.03)<br>[59.4,71.7] | 63.3 (0.02)<br>[59.0,67.6] | 59.0(0.02)<br>[54.9, 63.2] | 26.0 (0.02)<br>[20.9,31.0] | 25.0 (0.02)<br>[20.4,29.4] | 35.1 (0.02)<br>[31.4, 39.1] |
| **Midlife-onset decreasing**<br>**(n = 1,818)** | 23.3 (0.03)<br>[17.9,28.8] | 24.3 (0.02)<br>[21.0,27.6] | 25.3 (0.02)<br>[21.4, 29.2] | 63.0 (0.02)<br>[57.9,68.0] | 61.7 (0.02)<br>[57.0,65.4] | 58.7(0.02)<br>[55.2, 62.1] | 36.4 (0.03)<br>[31.1,41.8] | 32.1 (0.02)<br>[28.5,35.6] | 38.1 (0.02)<br>[34.4, 41.8] |
| **Stable-high symptoms**<br>**(n = 3,047)** | 50.0 (0.04)<br>[41.0,59.0] | 56.9 (0.02)<br>[52.6,61.2] | 59.3 (0.02)<br>[54.9, 63.6] | 45.3 (0.03)<br>[38.2,52.4] | 47.4 (0.03)<br>[42.1,52.8] | 43.1(0.02)<br>[38.8, 47.4] | 52.2 (0.03)<br>[45.5,59.0] | 45.8 (0.02)<br>[41.3,50.3] | 55.4 (0.02)<br>[50.9, 59.8] |
| **<u>BCS70 mental health trajectories</u>** |  |  |  |  |  |  |  |  |  |
| **Total**<br>**(n = 17,486)** | 23.1 (0.02)<br>[19.9,26.3] | 26.5 (0.01)<br>[24.8,28.4] | 27.6 (0.01)<br>[25.1,30.2] | 64.5 (0.01)<br>[61.7,67.4] | 60.6 (0.01)<br>[58.4,62.8] | 58.2 (0.01)<br>[55.9,60.6] | 27.8 (0.01)<br>[25.1,30.8] | 31.7 (0.01)<br>[29.1,34.3] | 32.4 (0.1)<br>[29.9,34.9] |
| **No symptoms**<br>**(n = 9,157)** | 5.5 (0.01)<br>[ 3.8,7.1] | 7.0 (0.02)<br>[5.8,8.2] | 8.7 (0.01)<br>[6.8,10.5] | 76.8 (0.02)<br>[73.7,79.9] | 75.2 (0.01)<br>[72.7,77.6] | 69.7 (0.01)<br>[67.2,72.2] | 16.0 (0.01)<br>[13.4,18.5] | 17.6 (0.01)<br>[15.3,20.0] | 19.4 (0.01)<br>[17.1,21.7] |
| **Early adult-onset decreasing**<br>**(n = 1,539)** | 18.4 (0.02)<br>[13.9,22.8] | 23.3 (0.02)<br>[19.9,26.7] | 24.6 (0.02)<br>[20.9,28.3] | 65.4 (0.03)<br>[61.2,69.5] | 62.4 (0.03)<br>[57.0,67.9] | 58.4 (0.02)<br>[54.6,62.2] | 31.2 (0.02)<br>[27.2,35.2] | 31.8 (0.02)<br>[27.6,35.9] | 34.5 (0.02)<br>[29.8,39.2] |
| **Adult-onset decreasing**<br>**(n = 1,902)** | 23.0 (0.03)<br>[16.6,29.4] | 27.4 (0.02)<br>[22.8,32.1] | 27.7 (0.03)<br>[22.4,33.0] | 63.4 (0.03)<br>[57.8,68.9] | 57.1 (0.03)<br>[51.9,62.3] | 55.0 (0.03)<br>[49.3,60.7] | 30.4 (0.03)<br>[25.1,35.8] | 34.8 (0.03)<br>[28.6,41.1] | 36.4 (0.02)<br>[31.6,41.2] |
| **Midlife-onset increasing**<br>**(n = 1,324)** | 44.4 (0.03)<br>[38.1,50.7] | 48.1 (0.02)<br>[43.7,52.4] | 49.8 (0.02)<br>[44.8,54.8] | 52.6 (0.03)<br>[47.1,58.1] | 46.7 (0.02)<br>[42.1, 51.2] | 46.6 (0.02)<br>[42.3,50.9] | 38.4 (0.02)<br>[34.2,42.7] | 42.5 (0.02)<br>[37.9,47.2] | 41.6 (0.02)<br>[37.0,46.3] |
| **Stable-high symptoms**<br>**(n = 3,564)** | 62.7 (0.04)<br>[54.0,71.4] | 70.4 (0.02)<br>[65.9,74.9] | 70.1 (0.02)<br>[65.0,75.1] | 37.8 (0.02)<br>[32.9,42.8] | 29.1 (0.02)<br>[24.7,33.6] | 34.3 (0.02)<br>[30.0,38.9] | 52.2 (0.03)<br>45.5, 59.0] | 62.6 (0.02)<br>[57.4,67.6] | 60.0 (0.03)<br>[53.9,66.1] |

Wave 1 (May 2020), Wave 2 (September to October 2020) and Wave 3 (February-March, 2021) (Percentage (%), Standard Error (SE) and 95% Confidence Interval (CI))

The largest group had few or no symptoms (‘no symptoms’, NCDS n=6,255 (40.9%); BCS70 n=9,157 (52.4%)). There was also a group with persistent severe symptoms (‘stable-high symptoms’, NCDS n=3047 (19.9%); BCS70 n=3,564 (20.4%)), and a group with adult onset and more favorable outcomes (‘adult onset decreasing’, NCDS=1658 (10.8%); BCS70=1902 (10.9%)). Both cohorts also had a group with severe symptoms developing in mid-life; however, in the NCDS the outcome was more positive (‘midlife-onset decreasing n=1818 (11.9%)) whereas in the BCS70 the symptoms were increasing (‘midlife-onset increasing’, n= 1324 (7.6%)). The final group in the NCDS was repeated minor symptoms (‘stable-low symptoms’, n=2513 (16.4%), while in the BCS70 early adult onset with more favorable outcomes (‘early adult-onset decreasing’ BCS70=1539 (8.8%)).

### Descriptive of mental health outcomes during the pandemic

During the first national lockdown in May 2020, overall, 17.5% and 23.1% (mean scores available in supplementary Table S5) of cohort members in the NCDS and BCS70 respectively were associated with high psychological distress, and 27.1% and 27.8% with loneliness, while the majority (67.4% in the NCDS and 64.5% in the BCS) had high levels of life satisfaction. This pattern was broadly similar throughout the course of the pandemic (when lockdown restrictions were lifted and during the third lockdown).

Nevertheless, different levels of mental health during the pandemic were related to pre-pandemic trajectories of psychological distress (by age of onset and symptom severity). 3.7% and 5.5% of individuals with no prior life-course psychological distress symptoms in the NCDS and BCS70 respectively, experienced high psychological distress early during the pandemic, compared to 9.4% to 50% in the NCDS and 18.4% to 62.7% in the BCS70 for other prior life course mental health groupings that had experienced pre-pandemic psychological distress symptoms at least once. Likewise, in both cohorts the proportion of individuals with feelings of loneliness and lower life satisfaction during the pandemic varied by trajectories of psychological distress; although the highest proportions were associated with ‘stable-high symptoms’, those with onset at different stages in adulthood and with varying levels of psychological distress prior to the pandemic were related to higher proportions of poorer mental health outcomes during the pandemic, than those with no prior symptoms.

### Risk of poor mental health during the pandemic by trajectories of psychological distress

Table 2 presents the relative risks in the fully adjusted models, associated with each of the mental health outcomes during the pandemic for different trajectories of psychological distress, with the largest group ‘no symptoms’ used as the reference category in the analysis (Additional analysis comparing alternative reference groups are available in supplementary Tables S7a and S7b).

**Table 2:** **Influence of psychological distress trajectories on the relative risk (RR) of mental health outcomes during the COVID19 pandemic**

|  | Psychological distress<br>RR (SE), [95% CIs] |  |  | Life satisfaction<br>RR (SE), [95% CIs] |  |  | Loneliness<br>RR (SE), [95% CIs] |  |  |
| --- | --- | --- | --- | --- | --- | --- | --- | --- | --- |
|  | Wave 1 | Wave 2 | Wave 3 | Wave 1 | Wave 2 | Wave 3 | Wave 1 | Wave 2 | Wave 3 |
| **<u>NCDS mental health trajectories</u>** |  |  |  |  |  |  |  |  |  |
| **Ref: No symptoms** |  |  |  |  |  |  |  |  |  |
| **Stable-low symptoms** | 2.40 (0.32)***<br>[1.85, 3.13] | 1.82 (0.23)***<br>[1.43, 2.32] | 2.14 (0.22)***<br>[1.75, 2.61] | 0.94 (0.02)*<br>[0.90, 0.99] | 0.95 (0.02)**<br>[0.91, 0.98] | 0.90 (0.02)***<br>[0.86, 0.94] | 1.46 (0.14)***<br>[1.21, 1.76] | 1.50 (0.12)***<br>[1.28, 1.75] | 1.54 (0.08)***<br>[1.38, 1.72] |
| **Adult-onset decreasing** | 4.20 (0.70)***<br>[3.04, 5.82] | 3.26 (0.41)***<br>[2.54, 4.18] | 3.69 (0.37)***<br>[3.03, 4.49] | 0.86 (0.03)***<br>[0.80, 0.92] | 0.81 (0.03)***<br>[0.76, 0.86] | 0.82 (0.03)***<br>[0.76, 0.89] | 1.66 (0.19)***<br>[1.33, 2.08] | 1.82 (0.20)***<br>[1.47, 2.25] | 1.88 (0.11)***<br>[1.67, 2.10] |
| **Midlife-onset decreasing** | 5.80 (0.76)***<br>[4.49, 7.49] | 4.02 (0.48)***<br>[3.18, 5.09] | 4.19 (0.40)***<br>[3.46, 5.07] | 0.82 (0.03)***<br>[0.76, 0.88] | 0.78 (0.03)***<br>[0.73, 0.83] | 0.81 (0.03)***<br>[0.76, 0.87] | 2.43 (0.20)***<br>[2.07, 2.85] | 2.40 (0.19)***<br>[2.05, 2.81] | 2.15 (0.15)***<br>[1.87, 2.47] |
| **Stable-high symptoms** | 10.78 (1.34)***<br>[8.45, 13.74] | 8.28 (0.93)***<br>[6.64, 10.32] | 8.57 (0.71)***<br>[7.27, 10.10] | 0.60 (0.03)***<br>[0.54, 0.68] | 0.61 (0.03)***<br>[0.56, 0.67] | 0.61 (0.02)***<br>[0.56, 0.66] | 3.21 (0.22)***<br>[2.80, 3.68] | 3.24 (0.29)***<br>[2.71, 3.86] | 2.85 (0.15)***<br>[2.56, 3.17] |
| **N 15,291** |  |  |  |  |  |  |  |  |  |
| **<u>BCS70 mental health trajectories</u>** |  |  |  |  |  |  |  |  |  |
| **Ref: No symptoms** |  |  |  |  |  |  |  |  |  |
| **Early adult-onset decreasing** | 3.23 (0.41)***<br>[2.51, 4.15] | 3.20 (0.31)***<br>[2.64, 3.88] | 2.70 (0.28)***<br>[2.19, 3.32] | 0.86 (0.03)**<br>[0.81, 0.91] | 0.84 (0.04)***<br>[0.77, 0.91] | 0.85 (0.03)***<br>[0.80, 0.91] | 1.87 (0.15)***<br>[1.59, 2.19] | 1.79 (0.14)***<br>[1.53, 2.10] | 1.76 (0.12)***<br>[1.53, 2.02] |
| **Adult-onset decreasing** | 4.11 (0.68)***<br>[2.98, 5.68] | 3.82 (0.41)***<br>[3.10, 4.72] | 3.10 (0.31)***<br>[2.52, 3.80] | 0.83 (0.03)***<br>[0.76, 0.90] | 0.76 (0.03)***<br>[0.70, 0.83] | 0.79 (0.04)***<br>[0.72, 0.87] | 1.86 (0.17)***<br>[1.56, 2.22] | 1.94 (0.16)***<br>[1.65, 2.28] | 1.87 (0.15)***<br>[1.58, 2.20] |
| **Midlife-onset increasing** | 7.91 (1.11)***<br>[6.00, 10.41] | 6.68 (0.56)***<br>[5.66, 7.87] | 5.51 (0.55)***<br>[4.50, 6.75] | 0.68 (0.04)***<br>[0.62, 0.76] | 0.62 (0.03)***<br>[0.57, 0.69] | 0.68 (0.03)***<br>[0.61, 0.74] | 2.33 (0.19)***<br>[1.99, 2.72] | 2.38 (0.18)***<br>[2.05, 2.77] | 2.13 (0.15)***<br>[1.84, 2.46] |
| **Stable-high symptoms** | 10.82 (1.36)***<br>[8.45, 13.85] | 9.40 (0.86)***<br>[7.86, 11.24] | 7.45 (0.68)***<br>[6.19, 8.97] | 0.50 (0.03)***<br>[0.45, 0.56] | 0.40 (0.03)***<br>[0.35, 0.46] | 0.50 (0.03)***<br>[0.44, 0.56] | 3.06 (0.23)***<br>[2.64, 3.55] | 3.39 (0.20)***<br>[3.02, 3.81] | 3.04 (0.19)***<br>[2.68, 3.46] |
| **N: 17,486** |  |  |  |  |  |  |  |  |  |

Reported as Relative Risk (RR), standard error (SE), †p<.10, \*p<.05, \*\*p<.01, \*\*\*p<.001, 95% confidence intervals (95% CIs)
Wave 1 (May 2020), Wave 2 (September to October 2020) and Wave 3 (February-March 2021)

#### Psychological distress

In both cohorts, any pre-pandemic experience of psychological distress across the ‘adult’ life course irrespective of age of onset, severity, longevity and proximal occurrence were associated with greater relative risk of high psychological distress during the pandemic. Mental health classes associated with the greatest risk of high psychological distress during the pandemic, had more than one prior episode of poor mental health and more recent occurrences; for example during the first lockdown ‘stable-high symptoms’ were associated with a 10.8-fold increased risk in the NCDS [95%CI: 8.5, 13.7] and BCS70 [95%CI: 8.5, 13.9], and the ‘midlife-onset increasing’ group in the BCS70 was related to a 7.9 (6.0, 10.4) fold increase of high psychological distress. Furthermore, ‘early adult-onset decreasing’ (mid 20’s) in the BCS70 was associated with more than 3 [95%CI: 2.5, 4.2] times the risk, and ‘adult-onset decreasing’ (early 30’s) in the BCS70 was 4.1 [3.0, 5.7] and in the NCDS 4.2 [3.0, 5.8] times the risk of high psychological distress, despite all groups having more favorable mental health outcomes prior to the outbreak of COVID-19.

#### Life satisfaction

In both cohorts all psychological distress trajectories (compared to ‘no symptoms’) were associated with a reduction in life satisfaction during the pandemic. In the NCDS, the risk for a reduction in life satisfaction for the ‘stable-high symptoms’ class was 40% [0.54, 0.68], while for the BCS70 the risk was associated with a 50% [0.45, 0.56] reduction in life satisfaction during the first lockdown. In addition, in the BCS70 the likelihood of life satisfaction during the pandemic was lowered by a third (RR=0.68 [0.62, 0.76]) for the ‘midlife-onset increasing symptoms’ class.

#### Loneliness

In both cohorts all trajectories of prior psychological distress, (compared with ‘no symptoms’) were associated with an increased relative risk of feelings of loneliness. In particular, ‘stable-high symptoms’ were associated with over a three-fold risk (NCDS RR=3.2 [2.8, 3.7]; BCS70 RR=3.1 [2.6, 3.6]) of loneliness in both cohorts. Also, ‘midlife-onset decreasing’ in the NCDS (RR=2.4 [2.1, 2.9]) and ‘midlife-onset increasing’ in the BCS70 (RR=2.3 [2.0, 2.7]) were related with at least a doubling of the risk of feelings of loneliness.

The magnitude of the relative risk of mental health outcomes associated with each trajectory of psychological distress, was similar within cohorts at different stages.

### Life course psychological distress and mental health outcomes at different stages of the pandemic

Figure 2 A_F presents the predicted probability of high psychological distress -, life satisfaction and loneliness associated with psychological distress trajectories in the two cohorts at three time-points during the pandemic. In the NCDS, high psychological distress and life satisfaction were stable in each of the psychological distress trajectories from May 2020 to February/March 2021. However, in the NCDS there was an increase in loneliness for all groups (except the ‘midlife-onset decreasing’) from September/October 2020 when lockdown restrictions were relaxed, compared to February/March 2021 when the third lockdown was enforced. In the BCS70, for all the trajectories of psychological distress, the probability of worsening mental health outcomes during the pandemic was fairly stable across the three-time points.

**Figure 2:**
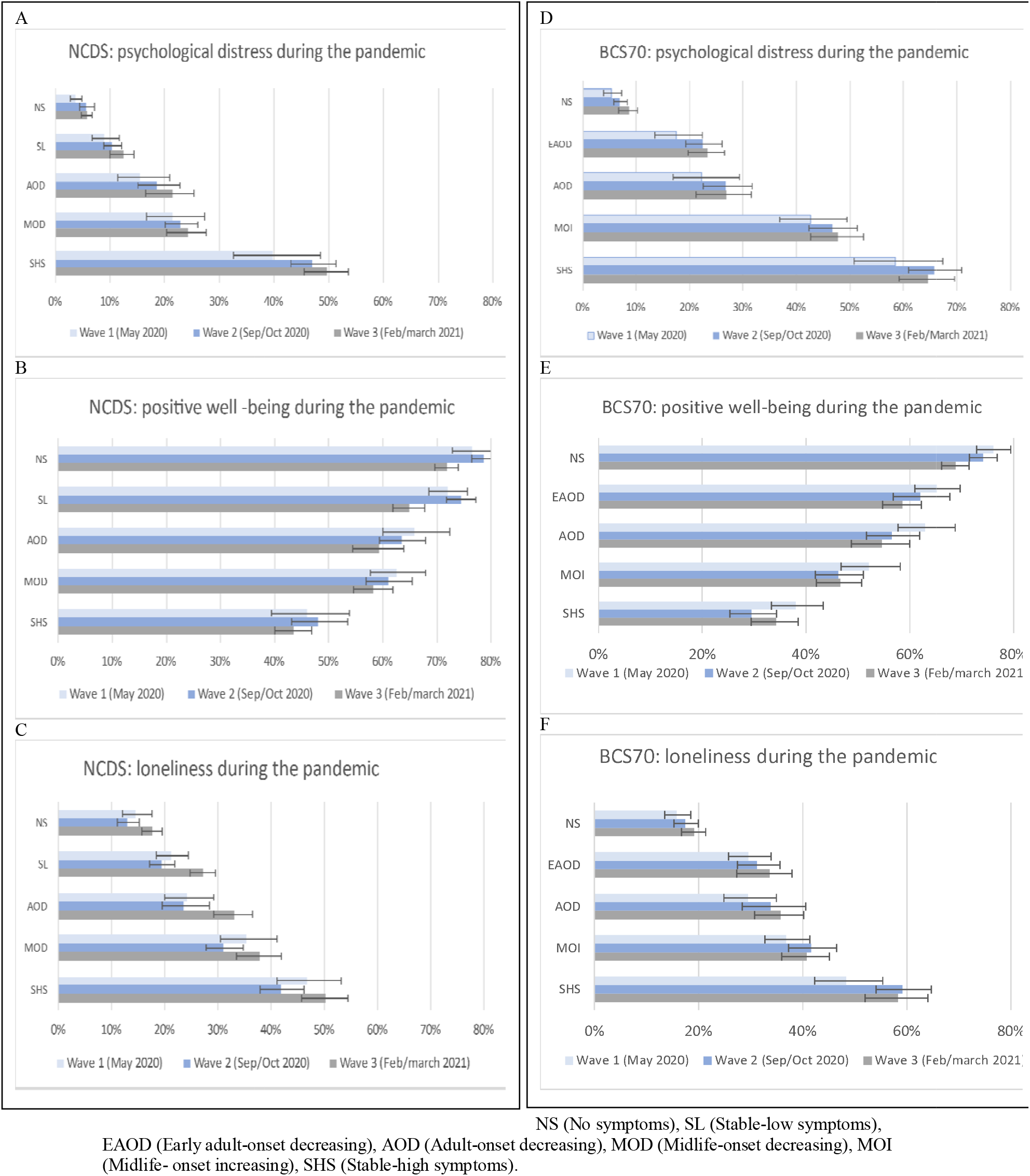
Predicted probability of mental health outcomes associated with psychological distress trajectories in the NCDS and BCS70 at different time-points during the pandemic.

## Discussion

We identified five distinct longitudinal typologies of psychological distress from adolescence to midlife. All groups with prior symptoms of psychological distress across the life course irrespective of age of onset, severity, longevity and proximal occurrence were associated with a greater relative risk of high psychological distress, reduction in life satisfaction, as well as feelings of loneliness during the pandemic. Pre-pandemic trajectories featuring more than one prior episode of poor mental health and more recent occurrences predicted the greatest risk of mental distress during the pandemic. The probability of high psychological distress, and lower life satisfaction (and loneliness for the BCS70) associated with psychological distress trajectories in the two cohorts remained fairly stable at three time-points during the pandemic, while in the NCDS, feelings of loneliness increased for most groups during the third lockdown when compared to early Autumn when restrictions had been relaxed.

Unlike other studies,^5^ which show a rapid decline and subsequent ‘bounce back’ in mental health, we show fairly stable mental health outcomes within trajectories of psychological distress during the pandemic. Indeed, a recent study^10^ shows the recovery of mental health did not occur in the UK until later (July 2020), coinciding with the lifting of lockdown measures, and during Autumn 2020, mental health deteriorated in particular groups as the pandemic developed. Our findings could reflect the distinct data collection timings; although the second data collection was during the lifting of lockdown measures in September/October 2020, the rule of six was introduced, the Prime Minister announced a second wave of coronavirus and the UK COVID-19 alert level moved from 3 to 4 (the epidemic is in general circulation, transmission is high or rising exponentially). The third data collection was conducted during the third national lockdown (amid rising hospitalization and death rates, and stay at home rules). However, other nationally representative studies have found life satisfaction and loneliness has remained largely stable throughout the pandemic.^1^

This study highlights the potential danger of dividing the population into symptomatic and asymptomatic groups. As other studies have illustrated symptoms are dimensional and can range from none to severe. In particular, the future mental health risks faced by those with subthreshold depression and anxiety have been shown to be associated with previous and future severe disorder. ^26, 27^ In addition, when comparing different pre-pandemic trajectories of psychological distress (Tables S7a and S7b), there were distinct relative risks associated with different mental health trajectories and mental health outcomes for most comparisons during the pandemic, thus illustrating the associated influence of heterogeneity of life-course mental health on future outcomes. Moreover, assuming mental well-being at one time point is indicative of an individuals’ predisposition to future mental health outcomes could underestimate the importance of life-course mental health for mental health outcomes during the pandemic.^6,7^ Population studies, focusing on depression have reported around 40-50% of those who recovered experienced a recurrence, ^28^ as well as comorbidity and transitions between disorders over time.^27^ In our study, the associated risk of poor mental health outcomes during the pandemic was greater for trajectories of psychological distress with recent occurrences, and with more than one prior episode. Indeed, those with prior mental health problems have been found to be at greater risk of post disaster mental health illness.^**Error! Reference source not found**.^

### Strengths and limitations

Our study is the first to relate longitudinal mental health data across more than 30 years before the COVID-19 pandemic within the same individuals to mental health outcomes during the COVID-19 pandemic. In addition, we identify groups of individuals with differing experience of psychological distress over time during adulthood in two cohorts, rather than treating longitudinal trajectories of mental health as homogenous. Other unique strengths include nationally representative samples, the sample size and prospective follow-up hitherto from birth to midlife, together with data collected at three discrete time points during the COVID-19 pandemic from May 2020 to March 2021.

There are a number of limitations of this study which are important to note. As with most longitudinal research, there was an issue of selective attrition. However, we included auxiliary variables in the multiple imputation models, including mental health and related variables from birth, which allows for predicting missing data with greater accuracy and minimizing non-random variation in these values.^29^ The study includes extensive data on pre-pandemic mental health. However, as we have no counterfactual at the time of the pandemic, we cannot assume the relative risks of poorer mental health outcomes were directly the result of the pandemic itself, ensuing circumstances and/or other factors. In terms of the method adopted, latent classes are approximations of symptom patterns in the data and do not represent actual data points, but are evidenced based summaries of mental health in the cohorts. In addition to this, with respect to the pre-COVID measures, we only had five ages where psychological distress was measured, and the most recent assessment was 4 and 12 years prior to the COVID-19 pandemic in the BCS70 and NCDS respectively. Furthermore, for both studies we used different measures and reporters (the parent at age 16 and self-report in adulthood) of psychological distress at age 16, compared to adulthood. We acknowledge that correlations between parent and self-report tend to be low.^30^ As self-report Malaise Inventory was available at age 16 in the BCS70 only, we reran the models and identified the same trajectories for the 5 classes identified using the original measures (Figure S1) and a difference in trajectory classification of only 2.9% (Table S8).

## Conclusions

Data from two British birth cohorts suggest that for different life-course trajectories of psychological distress there were distinct relative risks of poor mental health outcomes during the pandemic. Whilst any prior symptoms of psychological distress regardless of onset, severity and chronicity put individuals at greater risk, those with chronic and more recent occurrences were likely to require more support. We show fairly stable mental health outcomes during the pandemic for distinct life-course mental health groupings. Our findings show the importance of considering heterogenous mental health trajectories across the life-course in the general population in addition to mental health average population trajectories.

## Data Availability

Data is freely available from the UK Data Service at https://ukdataservice.ac.uk/

## References

1. Aknin L, De Neve JE, Dunn E, Fancourt D, Goldberg E, Helliwell JF, Jones SP, Karam E, Layard R, Lyubomirsky S, Rzepa A. Mental health during the First Year of the COVID-19 pandemic: a review and recommendations for moving forward. Perspectives on Psychological Science. 2021.

2. Daly M, Sutin AR, Robinson E. Longitudinal changes in mental health and the COVID-19 pandemic: Evidence from the UK Household Longitudinal Study. Psychological medicine. 2020; 1–10.

3. Pierce M, Hope H, Ford T, Hatch S, Hotopf M, John A, Kontopantelis E, Webb R, Wessely S, McManus S, Abel KM. Mental health before and during the COVID-19 pandemic: a longitudinal probability sample survey of the UK population. The Lancet Psychiatry. 2020;7(10):883–92.

4. Pan KY, Kok AA, Eikelenboom M, Horsfall M, Jörg F, Luteijn RA, Rhebergen D, van Oppen P, Giltay EJ, Penninx BW. The mental health impact of the COVID-19 pandemic on people with and without depressive, anxiety, or obsessive-compulsive disorders: a longitudinal study of three Dutch case-control cohorts. The Lancet Psychiatry. 2021;8(2):121–9

5. Fancourt D, Steptoe A, Bu F. Trajectories of anxiety and depressive symptoms during enforced isolation due to COVID-19 in England: a longitudinal observational study. The Lancet Psychiatry. 2021;8(2):141–9.

6. Kessler RC, Berglund P, Demler O, Jin R, Merikangas KR, Walters EE. Lifetime prevalence and age-of-onset distributions of DSM-IV disorders in the National Comorbidity Survey Replication. Archives of general psychiatry. 2005;62(6):593–602.

7. Moffitt TE, Caspi A, Taylor A, Kokaua J, Milne BJ, Polanczyk G, Poulton R. How common are common mental disorders? Evidence that lifetime prevalence rates are doubled by prospective versus retrospective ascertainment. Psychological medicine. 2010;40(6):899.

8. Colman I, Ploubidis GB, Wadsworth MEJ, Jones PB, Croudace TJ. A longitudinal typology of symptoms of depression and anxiety over the life course. Biological Psychiatry 2007; 62: 1265–1271.

9. Paksarian D, Cui L, Angst J, Ajdacic-Gross V, Rössler W, Merikangas KR. Latent trajectories of common mental health disorder risk across 3 decades of adulthood in a population-based cohort. JAMA psychiatry. 2016;73(10):1023–31.

10. Pierce M, McManus S, Hope H, Hotopf M, Ford T, Hatch SL, John A, Kontopantelis E, Webb RT, Wessely S, Abel KM. Mental health responses to the COVID-19 pandemic: a latent class trajectory analysis using longitudinal UK data. The Lancet Psychiatry. 2021 May 6.

11. Weich S, Brugha T, King M, McManus S, Bebbington P, Jenkins R, Cooper C, McBride O, Stewart-Brown S. Mental well-being and mental illness: findings from the Adult Psychiatric Morbidity Survey for England 2007. The British Journal of Psychiatry. 2011;199(1):23–8.

12. Frijters P, Clark AE, Krekel C, Layard R. A happy choice: wellbeing as the goal of government. Behavioural Public Policy. 2020; 4(2):126–65.

13. Santini ZI, Jose PE, Cornwell EY, Koyanagi A, Nielsen L, Hinrichsen C, Meilstrup C, Madsen KR, Koushede V. Social disconnectedness, perceived isolation, and symptoms of depression and anxiety among older Americans (NSHAP): a longitudinal mediation analysis. The Lancet Public Health. 2020;5(1):e62–70.

14. Luchetti M, Lee JH, Aschwanden D, Sesker A, Strickhouser JE, Terracciano A, Sutin AR. The trajectory of loneliness in response to COVID-19. American Psychologist. 2020;(75) 897–90

15. Power C, & Elliott J. Cohort profile: 1958 British birth cohort (national child development study). International journal of epidemiology 2006; 35: 34–41.

16. Elliott J, & Shepherd P. Cohort profile: 1970 British birth cohort (BCS70). International journal of epidemiology 2006; 35: 836–43.

17. Brown M, Goodman A, Peters A, Ploubidis G, Sanchez A, Silverwood R, Smith K. COVID-19 Survey in Five National Longitudinal Studies: Waves 1, 2 and 3: User Guide (Version 3). London: UCL Centre for Longitudinal Studies and MRC Unit for Lifelong Health and Ageing; 2021.

18. Little RJ, Rubin DB. Statistical analysis with missing data. John Wiley & Sons; 2019.

19. Mostafa T, Narayanan M, Pongiglione B, Dodgeon B, Goodman A, Silverwood RJ, Ploubidis GB. Missing at random assumption made more plausible: evidence from the 1958 British birth cohort. Journal of Clinical Epidemiology. 2021;136:44–54.

20. Rodgers B, Pickles A, Power C, Collishaw S, Maughan B. Validity of the Malaise Inventory in general population samples. Social psychiatry and psychiatric epidemiology. 1999;34(6):333–41.

21. McGee R, Williams S, Silva PA. An evaluation of the Malaise Inventory. Journal of Psychosomatic Research. 1986;30(2):147–52.

22. Furnham A, Cheng H. The stability and change of malaise scores over 27 years: Findings from a nationally representative sample. Personality and Individual Differences. 2015;79:30–4.

23. Cheung F, Lucas RE. Assessing the validity of single-item life satisfaction measures: Results from three large samples. Quality of Life research. 2014; 23(10):2809–18.

24. Steptoe A, Shankar A, Demakakos P, Wardle J. Social isolation, loneliness, and all-cause mortality in older men and women. Proceedings of the National Academy of Sciences. 2013;110(15):5797–801.

25. Pang M, Kaufman JS, Platt RW. Studying noncollapsibility of the odds ratio with marginal structural and logistic regression models. Statistical Methods in Medical Research. 2016 Oct;25(5):1925–37.

26. Bosman RC, Ten Have M, de Graaf R, Muntingh AD, van Balkom AJ, Batelaan NM. Prevalence and course of subthreshold anxiety disorder in the general population: a three-year follow-up study. Journal of affective disorders. 2019 Mar 15;247:105–13.

27. Merikangas KR, Zhang H, Avenevoli S, Acharyya S, Neuenschwander M, Angst J. Longitudinal trajectories of depression and anxiety in a prospective community study: the Zurich Cohort Study. Archives of general psychiatry. 2003 Oct 1;60(10):993–1000.

28. Mattisson C, Bogren M, Horstmann V, Munk-Jorgensen P, Nettelbladt P. The long-term course of depressive disorders in the Lundby Study. Psychological Medicine. 2007;37(6):883–92.

29. Sterne JA, White IR, Carlin JB, Spratt M, Royston P, Kenward MG, Wood AM, Carpenter JR. Multiple imputation for missing data in epidemiological and clinical research: potential and pitfalls. Bmj. 2009 29;338.

30. Collishaw S, Goodman R, Ford T, Rabe-Hesketh S, Pickles A. How far are associations between child, family and community factors and child psychopathology informant□specific and informant□general?. Journal of Child Psychology and Psychiatry. 2009, 50(5):571–80.

